# Determinants of quality of life in Latin American people with drug-resistant epilepsy: A cross-sectional, correlational study

**DOI:** 10.1101/2020.07.03.20146019

**Authors:** Marco Antonio Díaz-Torres, Edith Giselle Buzo-Jarquín, Aime Carolina Rodríguez-Martínez, Diana Laura De León-Altamira, Gerardo Padilla-Rivas, Sergio Andrés Castillo-Torres, Jaime Enrique Giovann Olivas-Reyes, J. Miguel Cisneros-Franco

**Author notes:** Address correspondence to: J. Miguel Cisneros-Franco, Montreal Neurological Institute, 753-3801 University, Montreal, QC, H3A 2B4, Canada, Jaime Enrique Giovann Olivas-Reyes, Servicio de Neurologia, Hospital, Universitario “Dr. Jose E. Gonzalez”., Universidad Autonoma de Nuevo Leon, Monterrey, NL, México. CP 64460.

## Abstract

One third of people with epilepsy (PWE) continue to have seizures despite adequate antiepileptic drug treatment. This condition, known as drug-resistant epilepsy (DRE) significantly impairs their social, family and work environment. The aims of this study were to assess the quality of life (QoL) in PWE with DRE and to investigate which factors are associated with a better QoL. This was a cross-sectional observational study of 133 Latin American PWE. QoL was assessed with the Spanish version of the Quality of Life with Epilepsy questionnaire (QOLIE-10). Independent clinical variables were analyzed with non-parametric statistics and their association with QoL was investigated with multiple linear regression. Poor quality of life was found in 25.8% of PWE. A low number of antiepileptic drugs (AEDs) was the major factor associated with better quality of life, closely followed by seizure frequency. We conclude that careful selection of AED treatment may contribute to improving both seizure control and QoL.

## Introduction

Epilepsy, one of the most frequent neurological diseases, is defined by the International League Against Epilepsy (ILAE) as “the enduring predisposition to suffer epileptic seizures” [1]. Drug resistance is found in 30% of people with epilepsy (PWE). This condition is associated with significant medical and psychiatric comorbidities, thus affecting the patient’s social, family and work environment.

The World Health Organization defines quality of life (QoL) as “the perception that an individual has about their position in life and in the context of their culture and value system in which they live in relation to their goals, expectations, standards and concerns” [2]. However, which factors are the main determinants of QoL in PWE remains unclear. Whereas some studies suggest that seizure frequency does not influence quality of life [3–5], others have identified drug-resistance as the main predictor of poor QoL [6], suggesting that seizure-freedom is necessary for optimal QoL [7–9]. Other studies, however, suggest that seizure severity, rather than seizure frequency, is a major determinant of QoL [10, 11]. Moreover, studies on the role of AED treatment in QoL have not produced consistent results [5, 10, 12].

We posit that these mixed results might be explained by idiosyncratic factors dependent on the specific cohort studied. And although previous studies have measured QoL in Latin American PWE [13–15], the specific determinants of QoL in patients with DRE remain understudied. To address this gap in knowledge, we set to study QoL in the outpatient clinic of our institution, a tertiary referral center with an overrepresentation of patients with DRE.

## Material and methods

### Patients and study design

This was a cross-sectional and observational study of 133 consecutive s PWE evaluated between the years 2016 and 2018, in the outpatient clinic of a tertiary academic center. The study included adult patients diagnosed with epilepsy for at least one year and according to the ILAE epilepsy diagnostic criteria [1]. Individuals with intellectual impairment, diseases that prevent the individual from understanding the questionnaire, those with a history of head trauma with massive bleeding, subjects with severe systemic diseases, non-epileptic seizures, and individuals with uncertain diagnosis or an incomplete medical record were excluded.

Clinical data, including classification of epilepsy, seizure type, seizure frequency, AED use, AED-associated adverse effects, etiology, age of epilepsy onset, and a depression diagnosis were obtained from the medical record. All patients were interviewed by a trained epileptologist, who verified the inclusion and exclusion criteria. The specialist also confirmed the diagnosis of epilepsy, detected cases of drug-resistant epilepsy according to ILAE criteria [16], and corroborated the syndromic diagnosis of each case. The clinical and demographic information was deposited in an institutional computer database for subsequent analysis.

### QoL assessment

The Quality of Life in Epilepsy-10 (QOLIE-10), Spanish version [17] was administered by a psychologist with experience in the field of neurology. The test comprises ten items rated from 1 to 5. It evaluates two components,; namely, Factor of Daily Activities and Impact of Treatment (DAIT, driving, social function, work, mental, physical and memory effects and mental health (overall score of quality of life, depression data, and the energy/fatigue ratio.) These values are given a score between 0 and 100, where 91–100 is excellent quality of life; 81–90 is very good; 71–80, good; 61–70, regular; and <60 poor.

The research subjects were grouped according to clinical and sociodemographic variables in order to compare the scores obtained in each component of the QOLIE-10: overall QoL, mental health, and DAIT.

### Statistical analysis

Analysis was done with SPSS version 23 (IBM). Normality was assessed with the Kolmogorov-Smirnov test. For continuous parametric variables, the mean is reported. Non-parametric variables regressed with clinical variables. Student’s t-test was used for parametric data, whereas Mann Whitney-U test and Kruskal-Wallis test were used for nonparametric data. Statistically significant variables (p < 0.05) were included in a linear regression model.

## Results

### Participants

A total of 133 individuals were included. The population was composed mainly of single individuals in their third decade of life with a basic education, and with focal and structural drug-resistant epilepsy (Table 1).

**Table 1.**
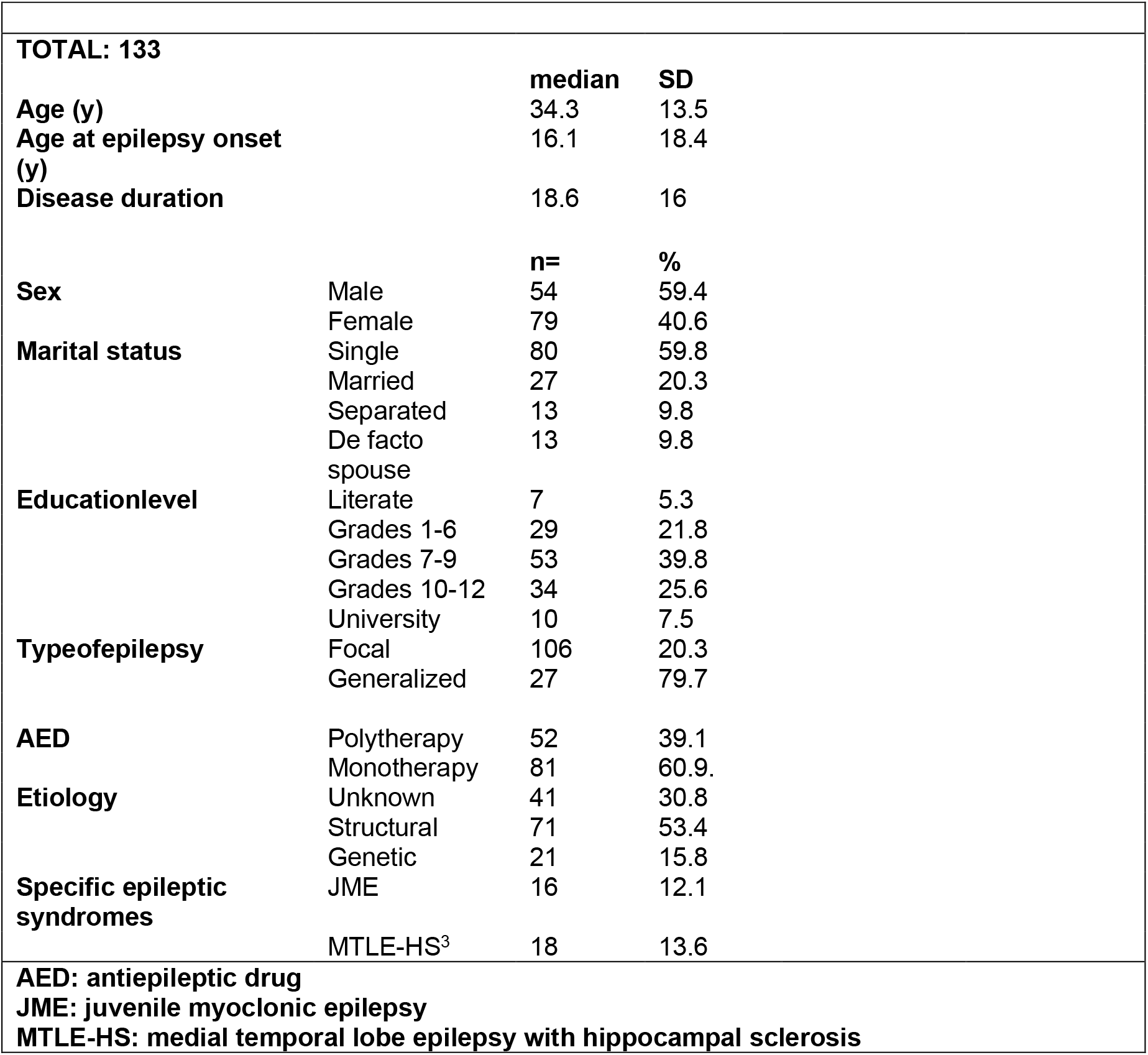
Clinical and demographic data

Valproate was the most widely used AED in polytherapy (42.5%) as well as in monotherapy (33.1%). Adverse drug events occurred in 34.5% of cases; the majority were non-serious adverse effects such weight gain or weight loss (17.4%, n = 23), mood alterations, 6.1% (n = 8), and drowsiness and occasional dizziness in 5.3% (n = 7). Other clinical and sociodemographic characteristics are summarized in Table 1.

Median (P50) total QOLIE score was 77.5 with an interquartile range of 60–87.5 and a range of 37.5–87.5 (Table 2).

**Table 2.** QOLIE-10 results

| Escalas | Poor<br>n (%) | Regular<br>n (%) | Good<br>n (%) | Verygood<br>n (%) | Excellent<br>n (%) |
| --- | --- | --- | --- | --- | --- |
| Mental Health | 45 (33.7) | 36 (27.2) | 16 (12) | 27 (20.3) | 9 (6.8) |
| Activities of<br>dailylife | 20 (21.8) | 15 (11.3) | 13 (9.8) | 25 (18.8) | 51 (38.3) |
| QoL | 34 (25.6) | 16 (12) | 32 (24.1) | 33 (24.8) | 18 (13.5) |

A majority of participants reported good, very good or excellent quality of life (62.4%, n = 83). Similarly, 66.9% of participants showed good, very good or excellent quality in activities of daily life and impact of antiepileptic treatment. In contrast, mental health was reported as “poor” in one third of our population (33.8%) (Table 3).

**Table 3.**
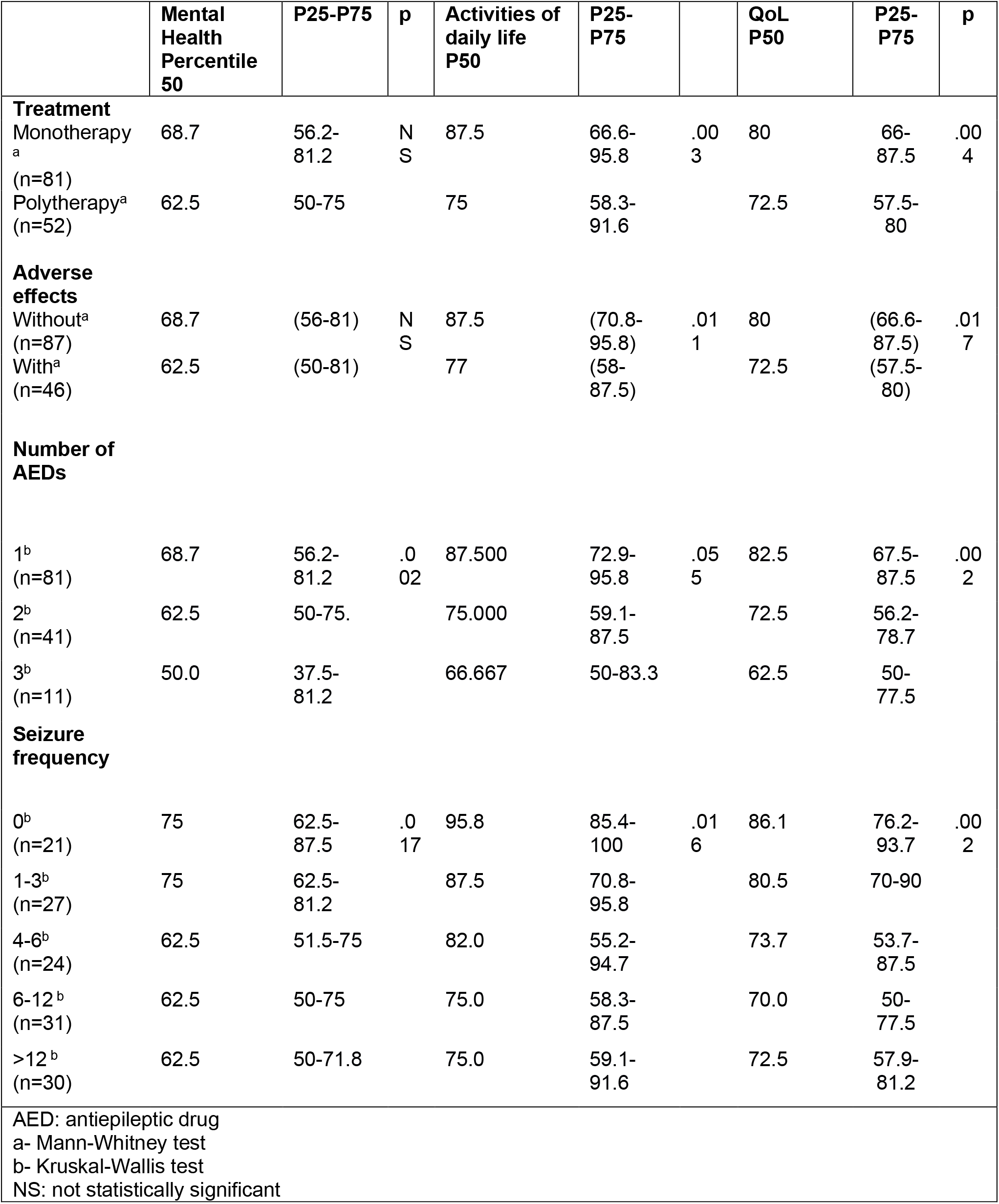
Differences in QoLfor PWE subgroups

|  | Mental Health Percentile 50 | P25-P75 | p | Activities of daily life P50 | P25-P75 |  | QoL P50 | P25-P75 | p |
| --- | --- | --- | --- | --- | --- | --- | --- | --- | --- |
| **Treatment** |  |  |  |  |  |  |  |  |  |
| Monotherapy<sup>a</sup><br>(n=81) | 68.7 | 56.2-81.2 | NS | 87.5 | 66.6-95.8 | .003 | 80 | 66-87.5 | .004 |
| Polytherapy<sup>a</sup><br>(n=52) | 62.5 | 50-75 |  | 75 | 58.3-91.6 |  | 72.5 | 57.5-80 |  |
| **Adverse effects** |  |  |  |  |  |  |  |  |  |
| Without<sup>a</sup><br>(n=87) | 68.7 | (56-81) | NS | 87.5 | (70.8-95.8) | .011 | 80 | (66.6-87.5) | .017 |
| With<sup>a</sup><br>(n=46) | 62.5 | (50-81) |  | 77 | (58-87.5) |  | 72.5 | (57.5-80) |  |
| **Number of AEDs** |  |  |  |  |  |  |  |  |  |
| 1<sup>b</sup><br>(n=81) | 68.7 | 56.2-81.2 | .002 | 87.500 | 72.9-95.8 | .055 | 82.5 | 67.5-87.5 | .002 |
| 2<sup>b</sup><br>(n=41) | 62.5 | 50-75. |  | 75.000 | 59.1-87.5 |  | 72.5 | 56.2-78.7 |  |
| 3<sup>b</sup><br>(n=11) | 50.0 | 37.5-81.2 |  | 66.667 | 50-83.3 |  | 62.5 | 50-77.5 |  |
| **Seizure frequency** |  |  |  |  |  |  |  |  |  |
| 0<sup>b</sup><br>(n=21) | 75 | 62.5-87.5 | .017 | 95.8 | 85.4-100 | .016 | 86.1 | 76.2-93.7 | .002 |
| 1-3<sup>b</sup><br>(n=27) | 75 | 62.5-81.2 |  | 87.5 | 70.8-95.8 |  | 80.5 | 70-90 |  |
| 4-6<sup>b</sup><br>(n=24) | 62.5 | 51.5-75 |  | 82.0 | 55.2-94.7 |  | 73.7 | 53.7-87.5 |  |
| 6-12<sup>b</sup><br>(n=31) | 62.5 | 50-75 |  | 75.0 | 58.3-87.5 |  | 70.0 | 50-77.5 |  |
| >12<sup>b</sup><br>(n=30) | 62.5 | 50-71.8 |  | 75.0 | 59.1-91.6 |  | 72.5 | 57.9-81.2 |  |
| AED: antiepileptic drug<br>a- Mann-Whitney test<br>b- Kruskal-Wallis test<br>NS: not statistically significant |  |  |  |  |  |  |  |  |  |

### Determinants of quality of life

To assess the relationship between QoL and a range of clinical and demographic variables, we compared QOLIE-10 scores between sub-groups of PWE determined as a function of these variables (Table 3).

Regarding pharmacological therapy, we found a significant difference in QoL as a function of the number of drugs used. In this population, those patients who used three or more antiepileptic drugs had lower overall QOLIE scores (median score: 1 AED, 82.5; 2 AEDs, 72.5; 3 or more AEDs, 62.5; p = .002; Table 3).

Patients medicated with three or more AEDs also had lower scores in the mental health component (MH median: 1 AED, 68.7; 2 AEDs, 62.5; 3 or more AEDs, 50; p = .002) and also in the component that evaluates daily activities and treatment impact (DATI median: 1 AED, 87.5; 2 AEDs, 75; 3 or more AEDs, 66.6; p = .055). Moreover, patients with slight adverse drug effects had lower daily activities and treatment impact scores (DATI median: with AE, 62.5; without AE, 68.7; p = .011), which were reflected as a worse overall quality of life (median QoL: with AE, 72.5; without AE, 80; p = 0.017) (Table 3).

We found no significant effect of the number or AEDs used on QoL sub-scores or in overall QoL. Nevertheless, we noted a trend whereby patients treated with more than two AEDs had lower overall QoL scores, regardless of the specific drugs used (MH median 50, p = .036; Table 3).

Patients with the highest number of seizures per year had lower scores in all of the QoL components (QL, p = .005; MH, p = .022; DATI, p = .026). Finally, no significant difference was found in QoL scores between patients with or without depression (28.4% and 31.6%, respectively).

### Linear regression

Building on the previous results, we included the variables number of seizures, number of current AEDs, and the variables listed in Table 4 in a linear regression model. We found that both seizure frequency (p = .010) and the number of current AEDs (p = .008) were significantly correlated with overall QoL. The beta coefficients indicated that both variables were inversely correlated with QoL, such that a greater number of drugs or greater seizure frequency were associated with greater work and social limitations, memory impairment, and lower MH scores (Table 4).

**Table 4.** Multivariate regression of QoL determinants

| Score and variables | Standardized B Coefficient | sig. | R<sup>2</sup> adjusted | 95% CI (lower, upperlimits) |  | Standardized B Coefficient | sig. | R<sup>2</sup> adjusted | 95% CI (lower, upperlimits) |  |
| --- | --- | --- | --- | --- | --- | --- | --- | --- | --- | --- |
|  | 1st regression |  |  |  |  | 2nd regression<br>Without non-significant variables |  |  |  |  |
| QoL |  |  |  |  |  |  |  |  |  |  |
| Adverse effects | -3.412 | N S | .098 |  |  |  |  | .109 |  |  |
| Seizure frequency | -3.253 | .007 |  | -5.109 | -.575 | -2.872 | .010 |  | -4.973 | -.614 |
| Number of AEDs | -5.951 | .018 |  | -10.27 | -.277 | -6.363 | .008 |  | -11.66 | -1.632 |
| Marital status | .720 | N S |  |  |  |  |  |  |  |  |
| Education | 1.495 | N S |  |  |  |  |  |  |  |  |
| Age at epilepsy onset | -.100 | N S |  |  |  |  |  |  |  |  |
| Etiology | -1.453 | N S |  |  |  |  |  |  |  |  |
| Depression | 3.892 | N S |  |  |  |  |  |  |  |  |
| Mental Health |  |  |  |  |  |  |  |  |  |  |
| Seizure frequency |  |  |  |  |  | -2.840 | .009 | .097 | -4.814 | -.670 |
| Number of AEDs |  |  |  |  |  | -5.278 | .023 |  | -9.843 | -.740 |
| Daily-life activities |  |  |  |  |  |  |  |  |  |  |
| Seizure frequency |  |  |  |  |  | -2.736 | .050 | .078 | -5.414 | .031 |
| Number of AEDs |  |  |  |  |  | -7.459 | .013 |  | -13.322 | -1.538 |
| NS- not statistically significant |  |  |  |  |  |  |  |  |  |  |

Finally, no significant differences were found in QoL as a function of sociodemographic factors such as gender, age, and educational level. Likewise, there were no significant differences in QoL as a function of clinical variables such as epilepsy type (focal vs. generalized epilepsy), time of evolution, seizure type, specific AED used (in case of monotherapy), type of epileptic syndromes, topographic diagnosis, etiology, and the presence of depression (Table 5).

**Table 5.** Variables not significantly associated with QoL (p-values)

| Scales | Mental Health | Daily-lifeactivities | QoL |
| --- | --- | --- | --- |
| Age<sup>a</sup> | .661 | .663 | .468 |
| Sex<sup>b</sup> | .138 | .744 | .554 |
| Education<sup>a</sup> | .209 | .670 | .557 |
| Age at epilepsy onset<sup>a</sup> | .861 | .721 | .803 |
| Duration of disease<sup>a</sup> | .174 | .256 | .217 |
| Epilepsy type<sup>b</sup> | .456 | .656 | .622 |
| Seizure type<sup>a</sup> | .691 | .128 | .298 |
| Etiology<sup>a</sup> | .314 | .622 | .341 |
| Syndromic diagnosis<sup>b</sup> | .482 | .551 | .422 |
| Topographic diagnosis<sup>a</sup> | .212 | .783 | .569 |
| Depression<sup>b</sup> | .873 | .839 | .795 |
| a-Kruskal-Wallis test |  |  |  |
| b-Mann Whitney test |  |  |  |

## Discussion

### Overall quality of life in drug-resistant epilepsy

Our work extends previous reports on PWE that, despite assessing QoL in different types of epilepsy, did not specifically study the effect of or made a distinction between the different degrees of control of each patient. In general, we found few impediments in daily activities and few social limitations despite the presence of drug-resistant epikepsy. QoL was good or excellent in 62.4% of cases. These results are at odds with two previous studies in Latin American PWE *without* DRE [15, 18]. These studies reported a higher proportion (43.3% and 31%, respectively) of PWE with a poor QoL (score < 60), relative to the present investigation (25.6%). However, in the perception of mental-emotional health we found similarities in comparison to other studies [18] where one third of patients stated difficulties in memory, energy, and low mood.

### Seizure frequency and pharmacological therapy

Multiple previous studies have concluded that, unless seizure freedom is achieved [7–9, 19], seizure frequency does not have a significant impact on quality of life [4, 5, 11, 20]. Our results, however, suggest that seizure frequency is a major determinant of QoL in people with DRE, as previously reported in studies in which individuals with higher seizure frequency exhibited a poorer quality of life [6, 14]. Although it was not possible to achieve complete control of seizures in patients with DRE, individuals with a lower seizure frequency had a better quality of life. These findings are in good agreement with previous studies in PWE with partial or total response to AED therapy [19–22] (but see [23]).

Antiepileptic therapy was another determinant factor of overall QoL, suggesting that, in terms of AEDs, “more does not always mean better”. Notably, our results suggest that, even in patients on polytherapy, fewer AEDs are associated with a better QoL [14].

The relationship between AED side effects of antiepileptic treatment and a poorer quality of life might be explained by two factors. First, impaired cognition associated with AED use may directly impact social interactions and other activities of daily life. Second, drug adverse effects may also potentiate the negative impact on quality of life caused by mood disorders (e.g., depression) and anxiety [21]. We therefore posit that an individualized approach, with special consideration on optimizing AED use, might translate into better social functioning, memory, and mood outcomes.

We theorize that antiepileptic polytherapy impairs the quality of life of patients by affecting social and functional aspects but also by concentration and memory impairments that affect the mental and emotional health of the individual and increasing the presence of side effects that considerably limit the functionality of the individual and his or her daily activities. Although this was not significant in this study, there is a possibility that the mere perception of adverse pharmacological effects could represent sufficient burden to impair quality of life [24]. In patients on polytherapy, determining which treatments are associated with better QoL outcomes represents a challenge, due to the existence of multiple combinations of AEDs.

It should be noted that the effect of AEDs on quality of life might be at least partially independent from that caused by the persistence of seizures. Although a greater number of drugs are often used in cases of patients with a greater number of seizures, these two factors may be considered independently. On the one hand a high frequency of seizures mainly affects independence and social function, increases the worry of suffering injuries and falls during seizures, and consequently affects quality of life [19–22]. On the other hand, the number of drugs might influence quality of life through adverse effects and drug interactions resulting from polypharmacy. This is consistent with studies such as those conducted by Auriel et al. [25] and Zou et al. [26], which single out adverse drug effects as the most important factor for quality of life in patients with freedom from seizures and who have suspended medication.

### Depression

Unlike studies by (Akdemir et al., 2016) and (Espinosa Jovel et al., 2016) that established a negative association between low quality of life, depression and etiology, we did not find significant differences in quality of life among patients with depression, nor between epilepsy of unknown, structural, and genetic etiology. However, in our study, the presence or absence of depression was determined from the clinical record and no depression screening scale was applied. Because of this there is a possibility that depression has been underdiagnosed.

### Limitations of the study

The study was conducted in a tertiary care academic hospital that provides health services to the patients across five neighboring states, most of which belong to low and middle socioeconomic status and who do not have access to other institutional medical services. For this reason, a significant proportion of our outpatient population consists of referred individuals with inadequate seizure control. Although the cost of medical visits for most patients (67.9% in our sample) is covered by the public health system, medications are not available on-site. For this reason, patients, or their relatives, must go to other institutions to fill their prescriptions. On a related note, the range of AEDs available through public drug insurance is limited to carbamazepine, phenobarbital, phenytoin and valproate. As a result, these drugs are widely used in our population even though there are better options with fewer side effects. Epilepsy surgery, although available in our region, is not covered by the public health system, thus making it more difficult to address drug resistance.

### Recommendations for future studies

Studies that plan to study the quality of life of patients with epilepsy should consider the role of psychiatric comorbidities such as anxiety and depression. These conditions are more frequent in patients with epilepsy and are related to a poor quality of life [4, 12, 27, 28]. However, as these diagnoses may be underreported in health records [4, 29, 30], we suggest the inclusion of a screening tool to detect these and other comorbidities. Subsequent studies should consider the perception that the individual has about their condition and how it relates to their social environment and health system [31].

## Conclusions

Despite mounting evidence that DRE has deleterious effects on multiple aspects of life in PWE, the practice of focusing solely on addressing seizure frequency and severity remains deeply rooted in current clinical practice. However, it is necessary to carefully choose the medication that will be most likely to achieve the dual goal of reducing seizure frequency with the fewest possible side effects.

In adequately selected cases of DRE, epilepsy surgery represents the best intervention to achieve freedom from seizures and AEDs, alongside improved quality of life [32]. However, this and other invasive therapy options remain unattainable for most drug- resistant patients in our clinical setting due to resource scarcity. This situation poses a dilemma in current clinical practice, because in order to improve QoL it is important to reduce the number of seizures; however, the addition of a second or even a third antiepileptic drug can negatively influence both the patient’s mental health and daily activities, resulting in a poor quality of life. Non-pharmacological therapies, including patient support groups, screening for anxiety and depression [33, 34], and counseling need to be offered as early as possible to improve social function and overall quality of life.

## Data Availability

Data is available upon reasonable request

## Abbreviations

PWE: people with epilepsy
AED: antiepileptic drug
QoL: quality of life
MH: mental health
DATI: daily activities and treatment impact
QOLIE: Quality of Life in Epilepsy questionnaire

## Author contributions

Conceptualization: M.A.D.-T.; Formal analysis: M.A.D.-T., J.E.G.O.-R., G.P.-R., E.G.B.- J.; Investigation: M.A.D.-T., J.E.G.O.-R., E.G.B.-J., A.C.R.-M., D.L.d.L.-A.; Project administration: M.A.D.-T., J.E.G.O.-R., A.C.R.-M., D.L.d.L.-A.; Supervision: M.A.D.-T.; Visualization: J.E.G.O.-R., G.P.-R.; Writing – original draft: J.E.G.O.-R., S.A.C.-T.; Writing – review & editing: M.A.D.-T., S.A.C.-T., G.P.-R., J.M.C.-F.

## Acknowledgements

The database for this study was created by Engineer José Luis González-Garza. Data entry was done in part by the Study Group Against Neurological Diseases (GECEN), Universidad Autónoma de Nuevo León School of Medicine (Monterrey, Mexico).

